# Trends in Utilization and Health Care Spending After Implementation of a Comprehensive Behavioral Health Program

**DOI:** 10.64898/2026.01.08.26343712

**Authors:** Graham Baum, Scott Graupensperger, Sara Khor, Casey Smolka, Millard Brown, Adam Chekroud, Matt Hawrilenko

## Abstract

**Objective:** To evaluate the impact of a comprehensive behavioral health (BH) program on healthcare utilization and total medical spending across 17 employer health plans.

**Study Setting and Design:** A retrospective evaluation of benefit implementation across 17 employer-sponsored health plans between November 1, 2019, and January 1, 2025. Interrupted time series analysis models were used to estimate shifts in utilization patterns and medical spending across the entire health plan population before and after program implementation, comparing observed trends (with the benefit) to counterfactual trends (without the benefit).

**Data Sources and Analytic Sample:** Medical claims data and program billing records from 17 health plans implementing the employer-sponsored benefit program, covering 854,579 employees and dependents. The analytic sample included all health plan enrollees to provide a generalizable estimate of benefit impact across the full health plan population, not limited to those who used the benefit.

**Principal Findings:** Program implementation was associated with a significant reduction in total medical spending across the full health plan population, inclusive of program costs. In program year 1, total medical spending decreased by 3.8% (95% CI, 0.01% to 7.51%), with further decreases in year 2 (8.9%; 95% CI, 2.47% to 15.28%), as a greater share of BH care was delivered through more cost-efficient and specialized outpatient services. While BH utilization initially increased following implementation, it gradually returned to expected levels, with 81% of program utilization in year 2 representing care that would have otherwise occurred through the traditional health plan. These patterns suggest the benefit facilitated earlier intervention in the care continuum and reallocated utilization toward more efficient services without increasing total system costs.

**Conclusions:** Expanding timely access to integrated behavioral health services can reduce total medical spending and shift care earlier in the continuum, particularly when delivered through a scalable, digitally-enabled platform. These findings support the use of integrated behavioral health programs as a prudent cost-containment strategy in employer-sponsored health plans.

## Introduction

Health care spending in the United States continues to rise at a pace that exceeds economic growth ^1^, placing growing financial pressure on employers who fund the majority of coverage through self-insured health plans. Behavioral health (BH) conditions are a major contributor to these costs as they are prevalent ^2,3^, frequently undertreated ^4^, and strongly associated with elevated downstream medical spending ^5–7^. In response, employers are increasingly adopting comprehensive BH benefit programs designed to expand timely access to evidence-based care. Although prior evaluations suggest these programs can improve clinical and financial outcomes among treatment-engaged individuals ^8,9^, the extent to which these effects persist over time or impact system-wide patterns of care across the full health plan population remains poorly understood.

BH has emerged as a key driver of overall health spending, with direct treatment costs increasing 12.3% annually from 2019 to 2022 ^10^, outpacing overall health spending growth ^11,12^. This rapid rise in demand for BH care has outstripped system capacity, contributing to persistent access gaps where only one in four individuals with a BH diagnosis receive care from a mental health professional within a year ^4^. These constraints amplify indirect costs, as undertreated BH conditions are associated with substantially higher medical spending due to prolonged symptom burden, complications with co-occurring physical conditions, and preventable hospitalizations ^5–7^. These substantial indirect costs underscore the need to evaluate how implementing BH benefit programs can expand utilization and shape spending trends over time across the broader health care system.

Understanding the temporal dynamics of launching a BH benefit is essential because the potential value of integrated BH care lies not simply in *whether* it reduces spending, but in *when* and *how* it shifts patterns of care ^13^. Early increases in BH utilization may reflect the program’s ability to absorb and reallocate care that would have otherwise emerged later through higher-cost channels. By facilitating earlier engagement and diverting individuals from more intensive or reactive services, integrated BH care can generate downstream reductions in medical spending with cost savings that typically accumulate gradually ^8,9,13^. Longitudinal medical claims data can provide unique insights into how utilization patterns evolve over time, when spending reductions occur, and whether cost offsets are temporary or sustained. These insights have direct implications for budgeting, forecasting, and evaluating the long-term return on investment in workforce mental health.

Although prior benefit evaluations offer insights into costs among individuals who use these programs ^8,9,14^, they provide limited visibility into how implementation affects broader spending trajectories across the whole healthcare system. Critically, without modeling underlying secular trends, it is difficult to determine whether observed changes reflect the program itself or ongoing shifts in healthcare utilization.

Understanding the broader impact of integrated BH care therefore requires analytic approaches that capture not only whether care utilization and spending changes, but how those changes evolve over time relative to expected, pre-existing trends.

### Current Study

To address these gaps, this study provides the first longitudinal, population-level evaluation of a comprehensive BH benefit implemented across 17 employer-sponsored health plans. Using full medical claims for more than 850,000 covered individuals, we assessed how benefit implementation influenced total medical spending and patterns of BH care across entire health plan populations. We employed an interrupted time series (ITS) design, a quasi-experimental approach suited for evaluating policy and program implementation in real-world settings ^15,16^. By modeling outcome trajectories before and after each employer’s launch date—and by explicitly accounting for underlying secular trends—the ITS design estimates the causal effect of the program on utilization and spending over time. This design supports a population-level analysis of system-wide changes in utilization and spending, capturing not only whether outcomes changed, but also when and to what extent those changes unfolded across the broader care delivery system. By examining the timing and magnitude of shifts in both BH and physical health spending, this study offers a generalizable assessment of how integrated BH care can alter care utilization patterns and function as a cost-containment strategy at scale.

## Methods

### Study design

This retrospective study used a quasi-experimental ITS design to evaluate the impact of implementing a comprehensive behavioral health benefit program across 17 employers. The study population included all employees and dependents enrolled in the employer-sponsored health plan for at least one month during the study period (November 1, 2019–January 1, 2025), totaling 854,579 individuals. By including the entire eligible population with no exclusion criteria, this study represents a full implementation design, capturing longitudinal changes in behavioral health utilization and medical spending across the health plan before and after benefit implementation.

Benefit launch dates varied by employer and marked the period where employees and dependents became eligible for the behavioral health benefit program. Employer launch dates and characteristics are provided in **Supplementary Table 1**.

Human subjects oversight was provided by the Yale University Institutional Review Board, which deemed this study “not human subjects” research and exempt from the need for informed consent (IRB protocol ID: 2000029276).

### Benefit Program

The study analyzed data from an employer-sponsored behavioral health benefit (Spring Health; Spring Care Inc), which has been demonstrated to improve clinical outcomes for depression and anxiety ^17,18^ and financial outcomes related to workplace productivity and retention ^8,9,14^. The program was designed to increase accessibility to care with an online platform providing personalized therapist recommendations based on patient needs, and rapid scheduling of video or in-person care, with the first available appointment available within 1.2 days on average ^18^. The program used evidence-based components (e.g., dialectical and cognitive-behavioral therapy), including measurement-based care with ongoing clinical assessments, unlimited care navigation sessions, and up to 12 free sessions with a mental health provider, up to 2 of which could be used for medication evaluation and treatment. Care beyond the employer-sponsored session limit was available as an in-network health plan benefit. Participation in the program was voluntary for all eligible individuals.

The program delivers care through a centralized data system, which integrates clinical documentation, assessment data, appointment scheduling, and billing. This centralized electronic health record (EHR) system enables rapid scheduling and measurement-based care with continuous monitoring of clinical outcomes, which standardizes the use of clinical assessments to guide treatment decisions and improve outcomes.

### Data Sources

*Medical claims data* included all medical claims incurred during the 12 months before program launch and 11-32 months after launch (mean=20.8, SD=8.0; see **Supplementary Table 1**).

*Allowed amounts*, representing the maximum amount the health plan will pay for a service, were used to calculate medical spending. Two employers only provided paid amounts, which were converted to allowed amounts using the following formula: Paid = Allowed * 0.8 ^8,19^. Allowed amounts were truncated at $50,000 per claim and $100,000 per month.

*Benefit program cost data* were obtained from each employer’s billing records and reflected employer-level total monthly costs for psychotherapy and medication management. Cost data included both the direct cost of behavioral health visits and non-clinical costs for services such as manager training and development, manager consultations, critical incident response, and access to a digital self-help app.

### Measures

*Utilization rate* was defined as the proportion of eligible employees and dependents with at least one behavioral health session in a given employer-month during the study period.

Medical costs were calculated using allowed amounts for the following outcomes:

*Total Medical Cost* included all medical claims plus benefit program costs.

*Physical Health Cost* included medical claims from all non-behavioral health categories, defined as claims without a primary or secondary diagnosis code from F01–F99 in the International Statistical Classification of Diseases, Tenth Revision (ICD-10). This encompassed spending across all standard physical health domains, including primary care and internal medicine, specialty care (e.g., cardiology, orthopedics), surgical procedures and hospitalizations, emergency services not coded for behavioral health, diagnostic imaging and laboratory tests, and physical therapy and ancillary services.

*Total Therapy Cost* included psychotherapy claims paid through the health plan, plus benefit program costs, but excluded all non-therapy claims.

*Total Behavioral Health Cost* included total therapy costs + any non-therapy claims with a mental health diagnosis code (International Statistical Classification of Diseases, Tenth Revision (ICD-10) code between F01 and F99).

*Non-therapy Behavioral Health Cost* included any inpatient, outpatient, emergency, or ancillary medical claims with a behavioral health diagnosis (ICD-10 F01–F99), excluding claims coded as psychotherapy or counseling sessions.

Time variables were defined as follows:

*Calendar months* represented the secular time trend, and were defined as the number of months from the midpoint of the study period (January 1, 2022).

*Program months* were included to capture the change in the trend after program implementation. They were operationalized as the number of months from the benefit launch date for each employer, with all pre-launch months coded as 0.

*Post-launch* was included to capture any immediate shift in utilization or spending after program implementation. It was included as a binary indicator of whether each employer-month was before the benefit launch (coded as 0) or after the benefit launch (coded as 1).

### Statistical Analysis

Interrupted time-series (ITS) models ^15,16^ were estimated using generalized linear mixed-effects regression to evaluate the impact of implementing the benefit program.

This approach leverages variation in program launch timing across 17 employers over a 5-year observation window. Each employer contributed monthly data before and after its implementation date, allowing us to model both within-employer changes and overall population-level trends. These data were aggregated to estimate a common time trend across calendar-months, representing the counterfactual or “Without Benefit” condition—that is, the expected outcome trajectory had the program not been implemented.

To estimate the program’s effect, we included a program-months variable coded as 0 for pre-implementation months and increasing by one for each month following launch. This term captures deviations from the counterfactual trend attributable to the benefit program, effectively quantifying the change in slope or level of spending trajectories post-implementation. The regression models accommodated different types of outcomes through linking functions: an identity link for continuous outcomes (e.g., medical costs), and a logit link for binary outcomes (e.g., utilization rate, modeled as a proportion). All models included a random intercept for employer to account for employer-level clustering of outcomes (e.g., those with different plan designs may have higher or lower overall spending). This approach ensures overall trend estimates are not biased by employers with higher or lower average spending levels. All models were fitted on employer-month aggregated data. Observations in each model were weighted by the total number of eligible individuals per employer-month. For utilization models, a binomial GLMM was specified where the outcome was the proportion of eligible individuals utilizing therapy or medication management services each month. Time variables (calendar months and program months) were modeled using natural cubic splines to flexibly capture nonlinear trends in the pre- and post-intervention periods. Calendar months were centered at the mid-point of the study period (January 1, 2022), providing approximately two years of pre- and post-launch observations.

Model selection was guided by fit metrics, including the Akaike Information Criterion (AIC), and robustness was evaluated through sensitivity analyses using alternative spline specifications. This process identified cubic spline models as the optimal functional form for utilization outcomes and linear specifications as the best fit for cost outcomes.

## Results

### Behavioral Health Utilization

Difference-in-differences results for behavioral health utilization are shown in **Figure 1** and **Supplementary Table 2**. Prior to benefit implementation, BH utilization through traditional health plans increased from 2.69% to 5.29% over four years, corresponding to a 19.6% year-over-year increase (95% CI, 15.0% to 24.2%; *p* < 0.001).

**Figure 1.**
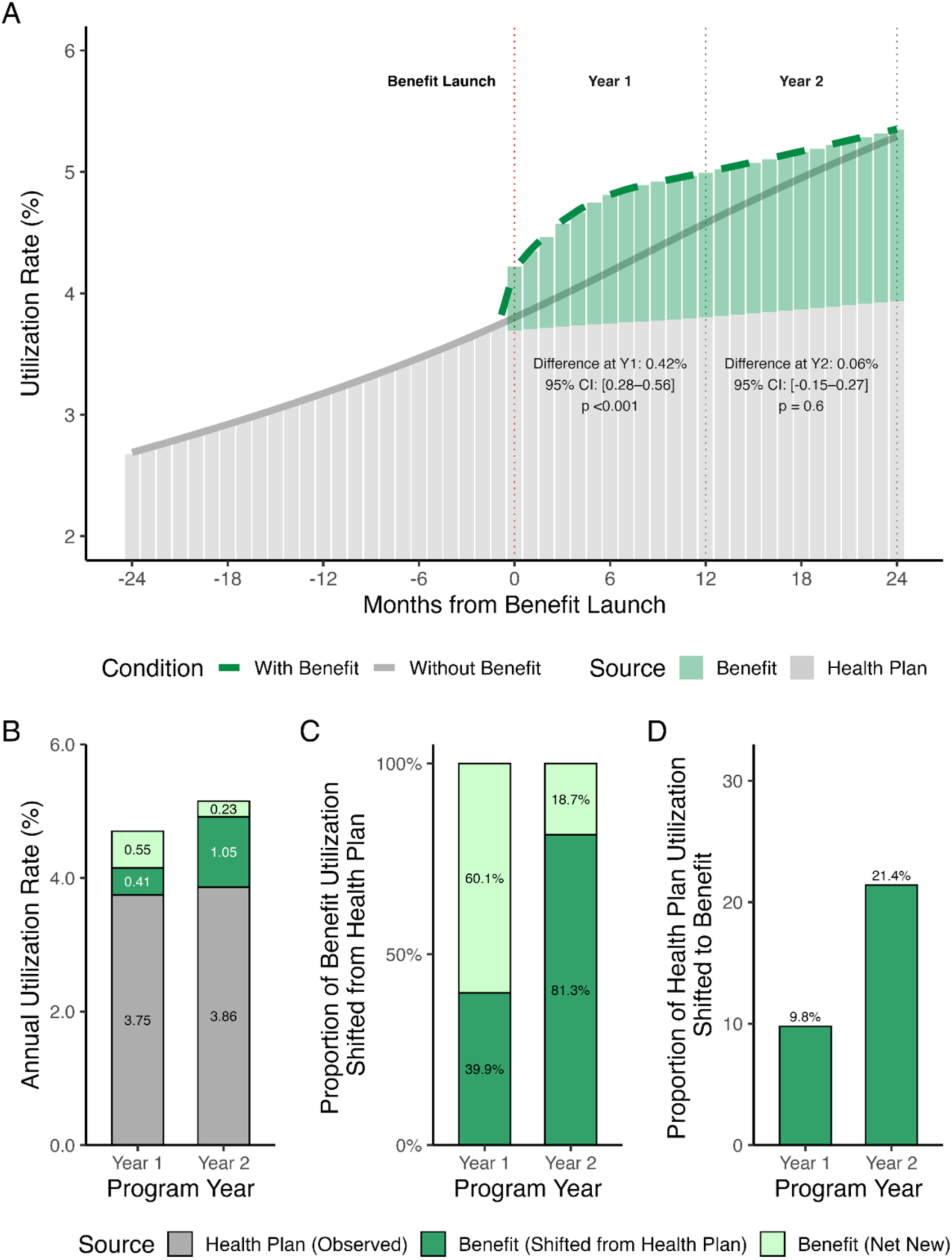
Behavioral health utilization following benefit implementation. (**A**) Benefit implementation (dashed green line) was associated with a significant immediate increase in behavioral health utilization relative to the health plan (solid gray line), followed by a flattening of the upwards trend. Shaded gray and green bars indicate utilization through the health plan and benefit program, respectively. The green bars above the gray line represent net new utilization — care that would not have occurred in that month without the benefit — while the green bars below the gray line represent care shifted from the health plan to the benefit. The sum of gray and green bars below the gray line represents the total utilization expected from the health plan in the absence of the benefit. (**B**) Overall, annual utilization increased over the first two program years, driven primarily by greater benefit use, which included both care shifted from the health plan (dark green) and net new care (light green). (**C**) The proportion of total benefit utilization that shifted from the health plan increased from 40% in year 1 to 81% in year 2, indicating reallocation of service delivery over time. (**D**) The proportion of expected health plan BH utilization that was delivered through the benefit increased from 10% to 21% over two years.

In the first month after benefit implementation, the overall utilization rate increased by 0.30 percentage points (*p* < 0.001) followed by a flattening of the trend. As shown in **Figure 1A**, total observed utilization (dashed green line) initially exceeded the counterfactual trend (solid gray line), then gradually converged with it by the end of year 2 (difference in month 24 = 0.06%, 95% CI, –0.15% to 0.27%; p = 0.60).

Relative to expected utilization, total behavioral health utilization was 13.2% higher in program year 1 (0.55% net new users / [0.41% shifted users + 3.75% health plan users]) and 4.7% higher in program year 2 (**Figure 1B**). In absolute terms, 0.96% of the total population used the benefit in year 1 and 1.28% in year 2.

As a share of total observed utilization, benefit-delivered care accounted for 20.4% of utilization in year 1 and 24.9% in year 2 (**Figure 1B**). Within benefit-delivered care, the proportion corresponding to care delivered through the benefit rather than the health plan increased from 39.9% in year 1 to 81.3% in year 2 (**Figure 1C**). Finally, relative to expected health plan utilization, the proportion delivered through the benefit increased from 10% in year 1 to 21% in year 2 (**Figure 1D**).

### Overall Medical Spending Trends

Implementing the benefit program was associated with a significant reduction in the upward trend of total monthly medical spending (**Figure 2A**). Prior to benefit launch, spend was trending upwards at a rate of $4.07 per person per month (PPPM). Following implementation, the monthly growth rate slowed to $1.77 PPPM, representing a reduction of $2.30 PPPM (95% CI, $-3.69 to $-0.92; *p* < 0.001; **Supplementary Table 3**).

**Figure 2.**
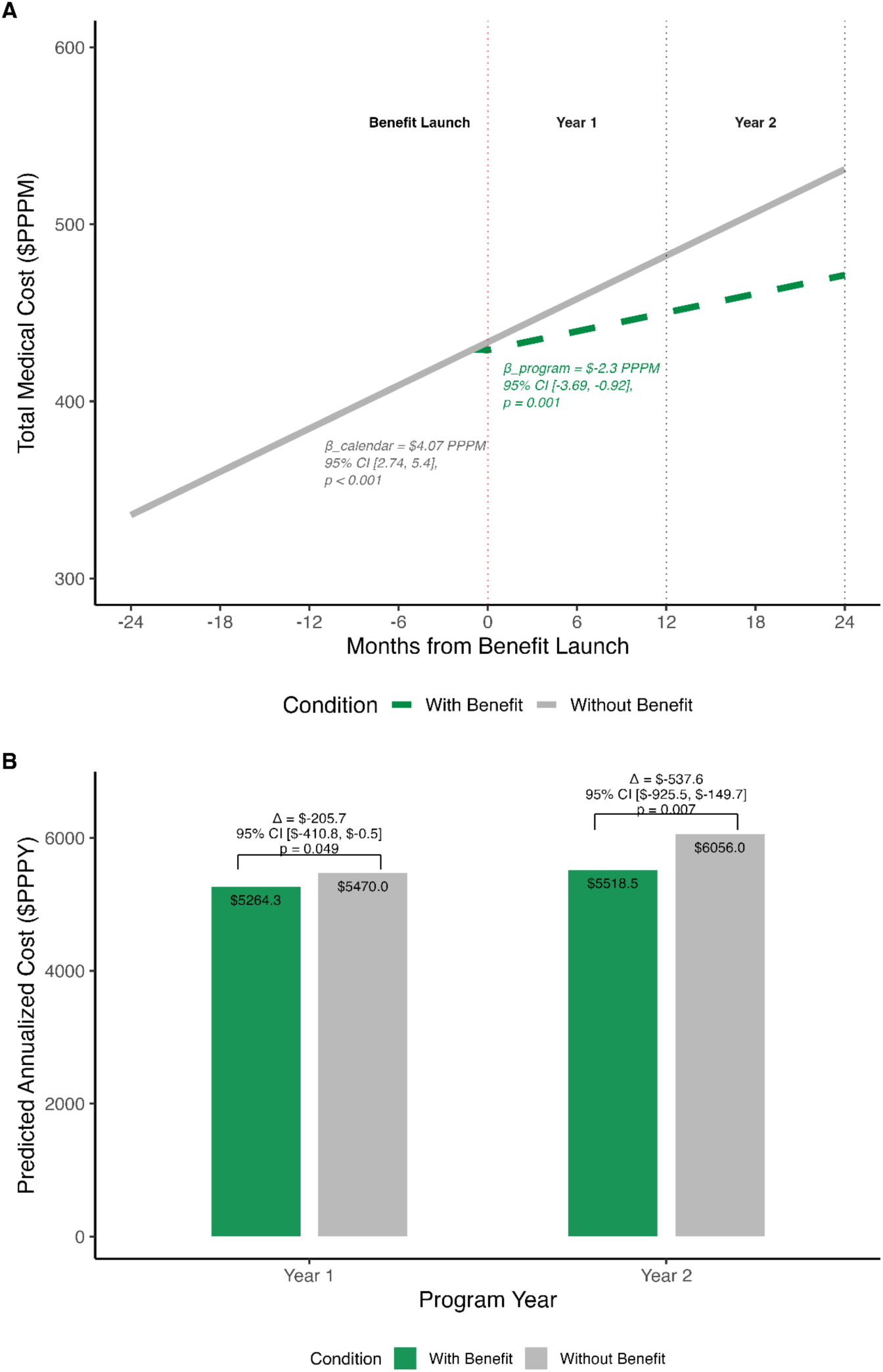
Total medical spending following benefit implementation. (**A**) Total monthly medical spending decreased significantly after benefit implementation. (**B**) Annualized cumulative spending per person per year (PPPY) after benefit implementation, reflecting a 3.8% relative reduction in year 1 and a 8.9% reduction in year 2.

As shown in **Figure 2B**, annualized cumulative medical spending was lower than expected in both program years. Annualized cumulative spending decreased by 3.8% in year 1 (β = $−205.65; 95% CI, $−410.77 to $−0.54; *p* = 0.049) and 8.9% in year 2 (β = $−537.57; 95% CI, $−925.47 to $−149.68; *p* = 0.007), relative to expected medical spending without of the benefit.

### Trend Breakdown by Physical and Behavioral Health

*Physical Health.* Reductions in total medical spending were driven primarily by changes in physical health spending. Prior to benefit implementation, physical health spending increased at a rate of $3.63 per person per month. Following implementation, the monthly growth rate slowed to $1.43 PPPM, corresponding to a $2.20 PPPM reduction in the rate of spending growth (95% CI, $-3.50 to $-0.91; *p* < 0.001; **Figure 3A** and **Supplementary Table 3**).

**Figure 3.**
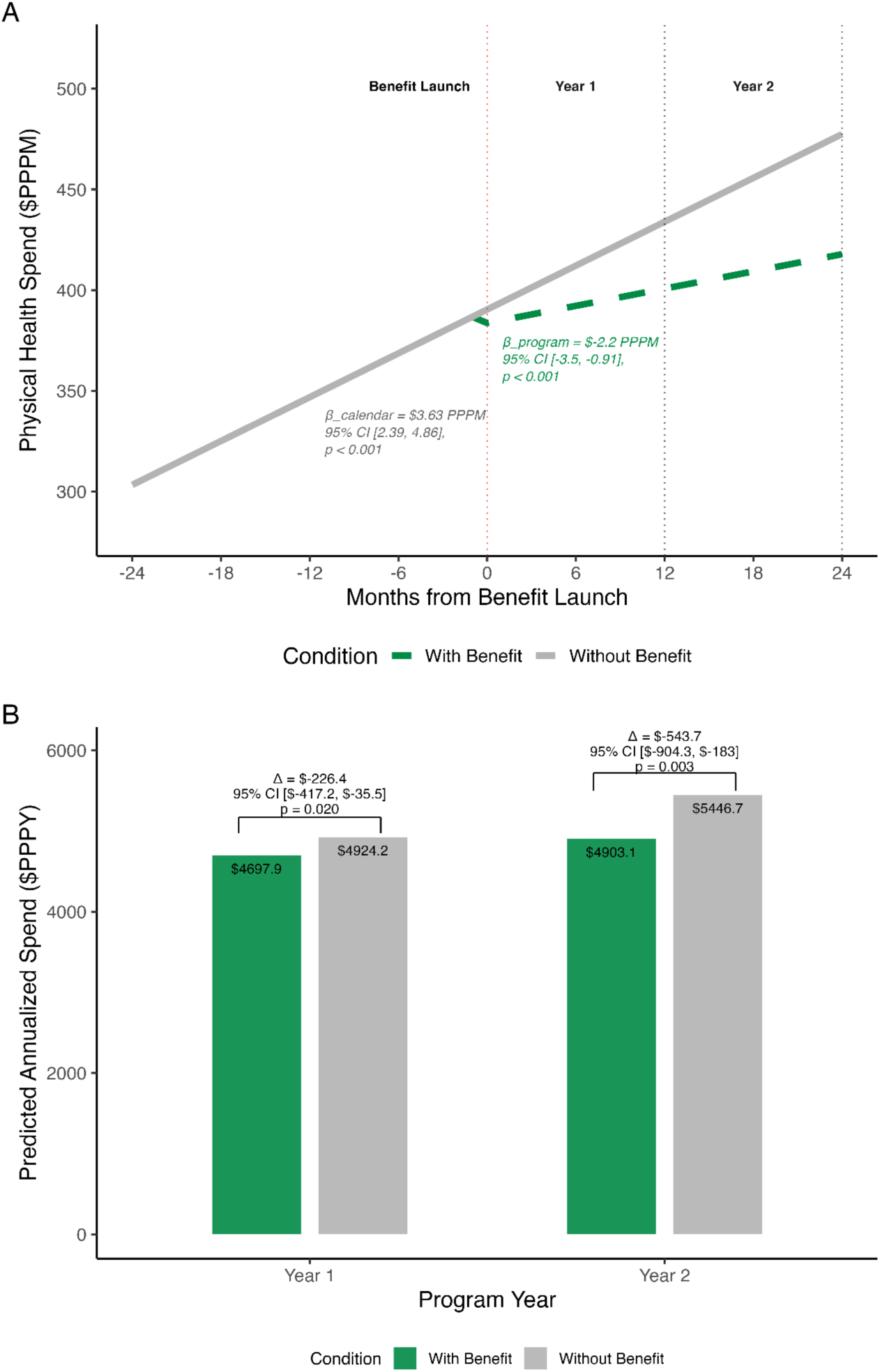
Physical health spending following benefit implementation. (**A**) Total monthly physical health spending decreased significantly after benefit implementation. (**B**) Annualized cumulative spending per person per year (PPPY) decreased over two years following benefit implementation, reflecting a 4.6% relative reduction in year 1 and a 10.0% reduction in year 2.

Consistent with these changes in monthly trends, annualized cumulative physical health spending decreased in both program years (**Figure 3B**), with reductions of $226.36 PPPY in year 1 (*p* = 0.02) and $543.65 PPPY in year 2 (*p* = 0.003).

*Behavioral Health.* Without the benefit, behavioral health spending increased at a rate of $0.43 PPPM (*p* < 0.001). Immediately following benefit implementation, total monthly behavioral health spending increased (βpost-launch = $2.40; *p* = 0.005; **Supplementary Table 3**). The post-implementation monthly trend did not differ significantly from the counterfactual trend (βprogram = -0.08; *p* = 0.45; **Figure 4A**).

**Figure 4.**
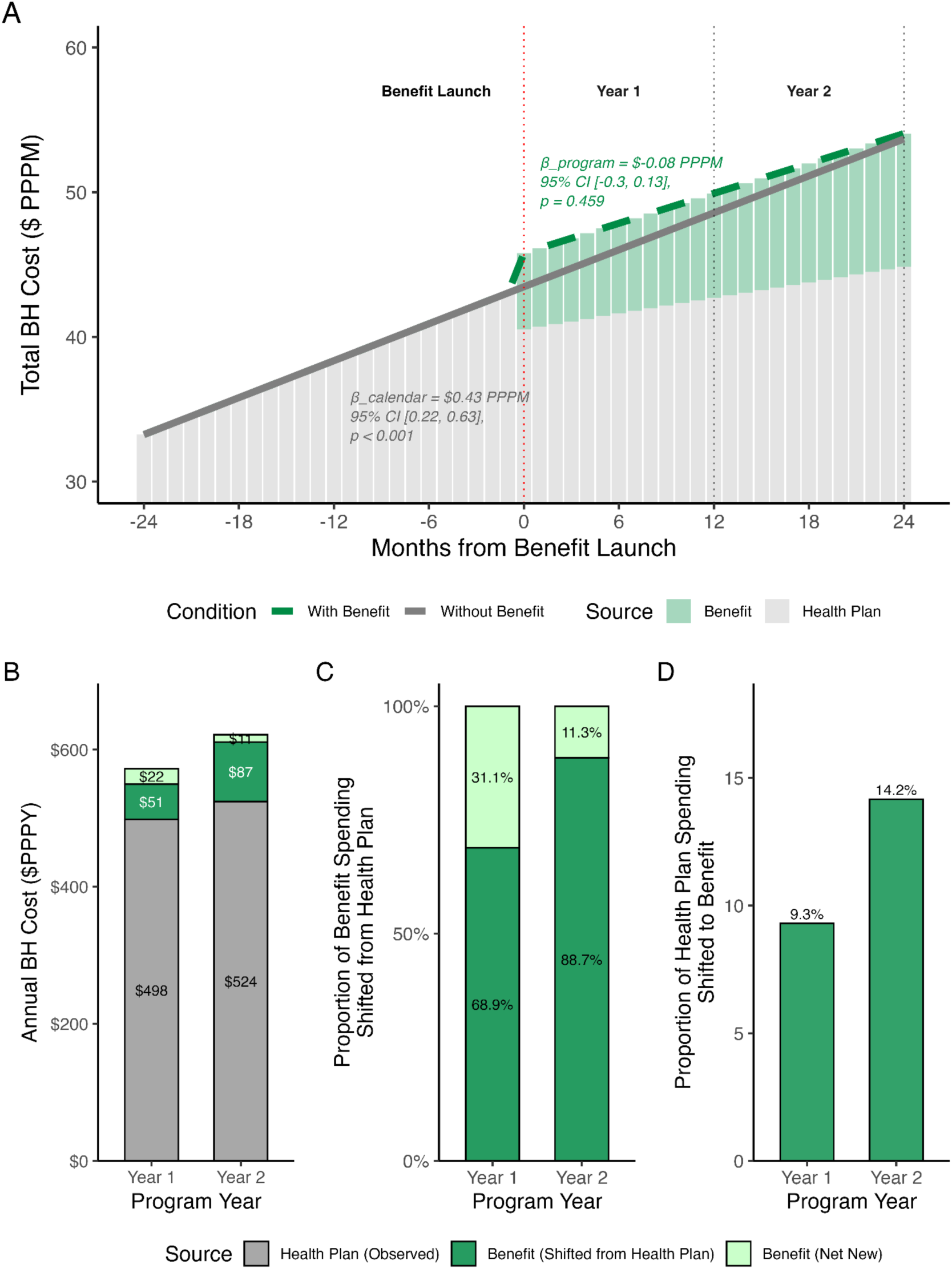
Behavioral health spending following benefit implementation. (**A**) Total monthly behavioral health costs had a modest level increase immediately after benefit implementation, but no significant change in the monthly trend. By the end of program year 2, observed spending with the benefit converged with expected spending through the health plan, indicating that program spending largely reallocated costs that would have occurred through the health plan rather than increasing total behavioral health spending. (**B**) Annual cumulative behavioral health spending increased over the first two program years, with a growing share delivered through the benefit program. Stacked bars show expected health plan spending (gray), benefit spending shifted from the health plan (dark green), and net new benefit spending (light green). (**C**) The proportion of total benefit spending that shifted from the health plan increased from 69% in year 1 to 89% in year 2, indicating a reallocation of behavioral health spending. (**D**) The proportion of expected health plan spending that was delivered through the benefit increased from 9% to 14% over two years.

Without the benefit, behavioral health spending increased by 11.8% per year (95% CI, 6.1% to 17.4%; *p* < 0.001). Following implementation, the benefit accounted for a growing share of total behavioral health spending —12.9% in year 1 and 15.7% in year 2 (**Figure 4B)**.

Within benefit-related spending, the proportion corresponding to spending delivered through the benefit rather than the health plan increased from 69% in year 1 to 89% in year 2 (**Figure 4C**). Correspondingly, the share of expected health plan BH spending delivered through the benefit increased from 9.3% to 14.2% in year 2 (**Figure 4D**).

### Therapy versus Other Behavioral Health Spending

*Therapy Spending.* Benefit implementation was associated with a significant immediate increase in therapy spending (βpost-launch = $2.82; *p* < 0.001; **Supplementary Table 3**). The monthly trend in therapy spending did not change significantly following implementation (βprogram = $0.05, *p* = 0.28), as shown in **Figures 5A** and **5B**.

**Figure 5.**
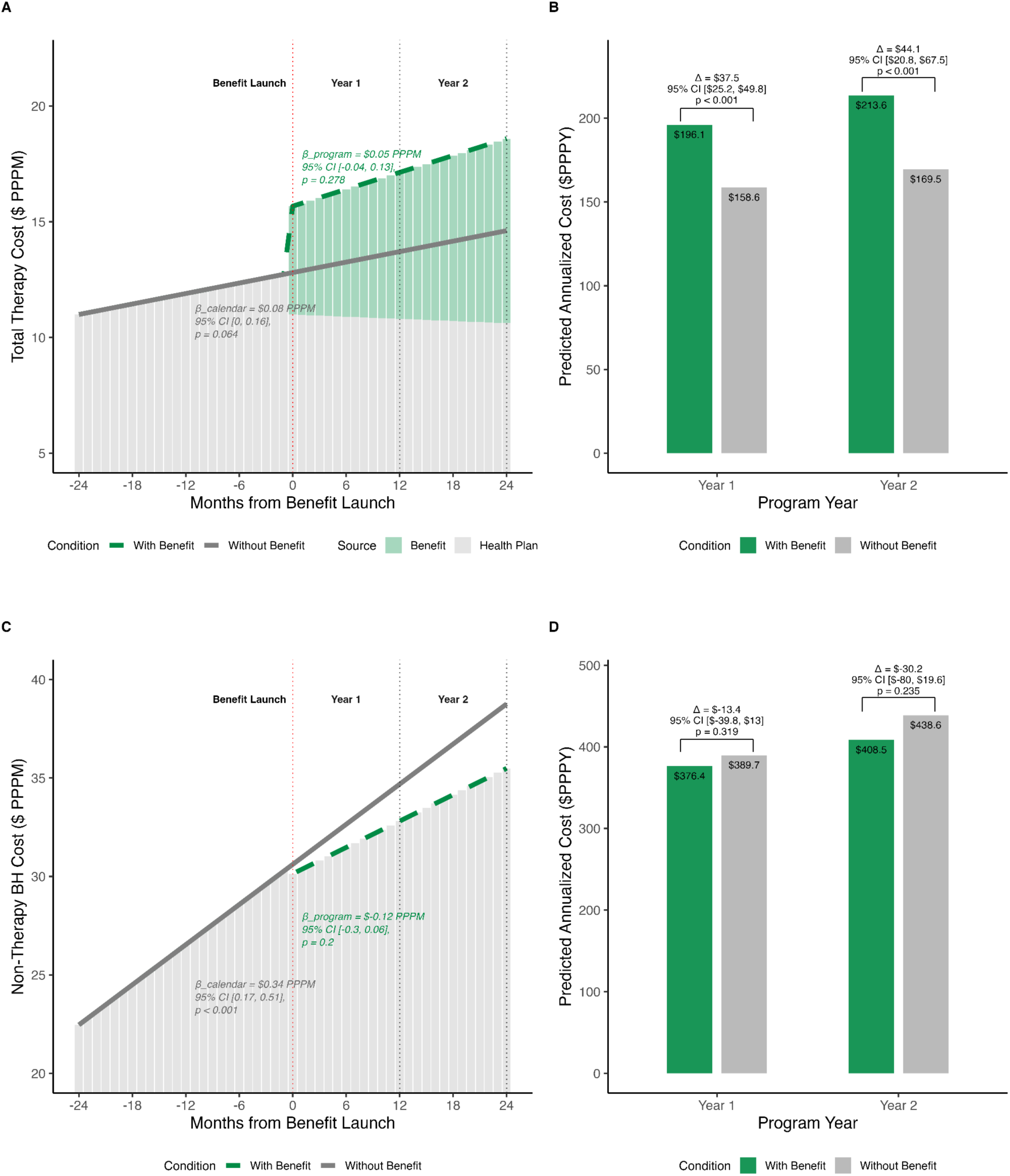
Therapy and non-therapy behavioral health spending following benefit implementation. (**A**) Monthly total therapy costs had a significant level increase immediately after benefit implementation, but no significant change in the rate of monthly spending. (**B**) Annualized cumulative therapy spending significantly increased with the benefit, with 23.6% higher spending in year 1 and 26.0% higher spending in year 2. (**C**) Monthly non-therapy behavioral health costs showed no significant immediate change in level or slope following benefit implementation. (**D**) Annualized cumulative spending decreased by 3.4% in year 1 and by 6.9% in year 2. Gray lines and bars represent the predicted counterfactual trend (Without Benefit), while green dashed lines and bars represent the observed trend (With Benefit). The vertical red dotted line in panels A and C indicates the timing of benefit launch.

*Non-Therapy Behavioral Health Spending.* Increases in therapy spending were partially offset by decreases in non-therapy behavioral health spending, comprising inpatient, emergency, and non-therapy outpatient visits (**Figure 5C**). Although neither the immediate level change (βpost-launch = $-0.36; *p* = 0.61) nor the change in monthly trend (βprogram = $–0.12; *p* = 0.20; **Supplementary Table 3**) were statistically significant, both contributed to modest reductions in cumulative spending over time. As shown in **Figure 5D**, annualized cumulative spending showed nonsignificant decreases of 3.4% in year 1 (β = $-13.39 PPPY, *p* = 0.32), and 6.9% in year 2 (β = $-30.19 PPPY; *p* = 0.23).

## Discussion

Employer-sponsored behavioral health benefits have become a cornerstone of healthy, high-functioning workplaces. However, evaluations have largely focused on program users relative to matched nonusers, offering limited insight into the system-level effects of these benefits, specifically when and how they shape utilization patterns and medical spending across the full health plan population. This study addresses that gap by conducting a population-level implementation analysis of a comprehensive BH benefit on utilization and total medical spending across 17 employer-sponsored health plans. In the first year after implementation, the benefit was associated with medical claims savings over and above the costs of the benefit program. Findings also highlighted temporal patterns of earlier entry into care, indicating that the benefit surfaced previously unmet behavioral health needs. Rather than simply shifting the timing of care, earlier access reduced the persistence and recurrence of need over time. Thus, the benefit program expanded access and treated more people without increasing total health care spending. These results provide evidence that expanding timely, integrated BH care can serve as a cost-containment strategy for employer-sponsored health plans.

Implementing the benefit initially expanded BH utilization, then consistently shifted care expected under the traditional health plan into the benefit. Overall, the benefit increased utilization by 13.2% relative to expectation in year 1, and 5.7% in year 2, reflecting the rapid identification and treatment of previously unmet behavioral health needs when care was made more easily accessible. Additionally, the benefit absorbed 9.8% of expected health plan utilization in year 1, and 21.4% in year 2, as the benefit increasingly served as the primary channel for meeting remaining behavioral health needs within a more efficient care delivery model. Importantly, by the end of year 2, combined health plan and benefit utilization converged with expected health plan utilization alone, suggesting that benefit use reduced the persistence and recurrence of care needs. This pattern is consistent with the program’s design—offering direct web scheduling, rapid access (median wait time 1.2 days^18^), and coordinated care delivery—which facilitates earlier engagement and increases the chance of successful resolution of behavioral health needs. Earlier engagement, in turn, may have reduced reliance on less coordinated and higher-cost health plan services.

The stabilization of overall utilization in year 2 further supports the interpretation that the program redistributed demand earlier in the care continuum. Several mechanisms may explain this pattern. First, rapid access and lowered barriers to care likely facilitated timely engagement for individuals with emerging but not yet acute needs. Second, the benefit’s centralized EHR, which enables measurement-based care and data-driven patient-provider matching ^20^, may have improved treatment efficiency by calibrating care to clinical need, increasing the likelihood of success in the first episode. This, in turn, reduces the need for additional care episodes, limits prolonged treatment courses, and prevents escalation to higher-cost services ^18,20,21^. Collectively, these dynamics suggest that comprehensive behavioral health programs can shift care to earlier, more efficient intervention points, stabilize longer-term utilization, and mitigate downstream cost escalation.

These utilization patterns were mirrored by shifts in BH spending over time. Notably, the benefit program expanded access to BH services without increasing total BH spending—costs rose modestly in year 1 (4.1%) and year 2 (1.8%) compared to expected spending without the benefit, but neither increase was statistically significant. By the second year, total BH spending converged with expected levels, with 89% of program spending representing costs that would have otherwise occurred through the health plan (**Figure 4C**). This normalization of spending reflects a transition from front-loaded treatment of unmet need to lower ongoing demand as patients completed courses of care and mental health conditions were managed or resolved. Previously reported shifts from higher-cost services, such as inpatient and emergency care, toward lower-cost outpatient therapy ^8^, are consistent with this interpretation.

Shifts in behavioral health spending were accompanied by growing reductions in total medical costs of 3.8% in year 1 and 8.9% in year 2, driven in part by reductions in physical health spending. Untreated or undertreated mental health conditions have long been associated with worse physical health ^22^ and increased physical health spending ^5,7,8,13,23–25^. The current data – showing the coupling of earlier behavioral health care (**Figure 1A**) with gradually accumulating reductions in physical health spending (**Figure 3**) – suggest a critical mechanism of care in which treating mental health conditions earlier in the continuum may contain subsequent increases in physical health spending. The continued growth of savings across program year 2 suggests that longer-term savings derive both from patients treated in year 1—who may no longer require active behavioral health treatment due to sustained improvement—and from new patients initiating care in year 2 whose needs are addressed earlier and more effectively. In this way, increasing the accessibility of quality behavioral health care may reduce both the incidence and duration of untreated behavioral health conditions, with compounding effects on medical spending.

This study advances our understanding of BH benefit programs by evaluating the system-level financial and utilization impact of program implementation across all health plan enrollees, not only those with a behavioral health diagnosis or those who engaged with the benefit. It is the first study to show the system-wide impacts of increasing accessibility to mental health care, and finds that many individuals using the benefit experienced earlier treatment that reduced ongoing behavioral health need and downstream physical health costs, rather than merely shifting care across settings. This design complements earlier research focused on treatment-engaged individuals using matched cohort designs ^8,9^, offering additional support for large-scale implementation of comprehensive behavioral health programs as a mechanism for reducing unmet need and preventing chronicity at the population level.

### Limitations

This study has several limitations. Although our modeling adjusted for pre-intervention trends, we cannot fully account for all concurrent changes in benefits or organizational dynamics that may have influenced outcomes. For example, employers that chose to adopt the benefit program may have done so because they were experiencing atypical cost trends, which could bias the counterfactual trend upwards. Similarly, the COVID-19 pandemic may have contributed to accelerated increases in behavioral health utilization and spending during the pre-launch period, potentially accounting for the steep counterfactual trend in behavioral health utilization found here. Relatedly, if employers launched other cost-containment strategies concurrently with the benefit program, it could bias program impacts upwards. However, the triangulation of current analyses centered on program launch dates with prior matched cohort studies centered on individual treatment dates supports the conclusion that the benefit was the primary driver of the observed differences.

## Conclusion

This study provides real-world evidence that increasing accessibility and utilization of behavioral healthcare can reduce total medical spending in employer-sponsored health plans. Critically, the benefit appeared to shift behavioral health spending earlier in the continuum and towards more efficient services, driving cost savings that increased over time. Together, these findings support the inclusion of integrated behavioral health programs as a viable strategy for improving access, delivering high-quality care, and containing healthcare costs across diverse employer settings.

## Data Availability

Due to the terms of our business agreements with the employers who shared medical claims data with us, we are not authorized to share individual-level data. Consequently, while we can provide aggregate findings and analyses, we are unable to share the raw data itself.

