## Supplementary Material for "Trends in Utilization and Health Care Spending After Implementation of a Comprehensive Behavioral Health Program"

| Employer | Start Date (Pre) | End Date (Pre) | Start Date (Post) | End Date (Post) | Employer Size (Pre) | Employer Size (Post) | BH Utilization Rate (Pre) | BH Utilization Rate (Post) | Months (Pre) | Months (Post) |
|---|---|---|---|---|---|---|---|---|---|---|
| Employer_1 | 2019-11-01 | 2020-10-01 | 2020-11-01 | 2021-10-01 | 5216 (639.4) | 6921 (481.2) | 5.8 (0.3) | 7.7 (0.3) | 12 | 12 |
| Employer_2 | 2020-02-01 | 2021-01-01 | 2021-02-01 | 2023-09-01 | 13936 (104.8) | 13950 (335.3) | 4.9 (0.7) | 7.9 (0.8) | 12 | 32 |
| Employer_3 | 2021-06-01 | 2022-05-01 | 2022-06-01 | 2023-05-01 | 11700 (134.9) | 11809 (68.7) | 6.2 (0.5) | 8.5 (0.5) | 12 | 12 |
| Employer_4 | 2022-01-01 | 2022-12-01 | 2023-01-01 | 2025-01-01 | 97212 (4544.4) | 104182 (1545.2) | 2.4 (0.1) | 3.1 (0.2) | 12 | 25 |
| Employer_5 | 2021-04-01 | 2021-12-01 | 2022-01-01 | 2024-01-01 | 11093 (406.3) | 12758 (470.1) | 5 (0.2) | 6.7 (0.4) | 9 | 25 |
| Employer_6 | 2021-01-01 | 2021-12-01 | 2022-01-01 | 2023-12-01 | 2854 (124.5) | 3154 (78.5) | 2.7 (0.2) | 3.4 (0.3) | 12 | 24 |
| Employer_7 | 2022-01-01 | 2022-12-01 | 2023-01-01 | 2024-03-01 | 13254(257.3) | 13833 (167) | 6 (0.3) | 8 (0.3) | 12 | 15 |
| Employer_8 | 2021-01-01 | 2021-09-01 | 2021-10-01 | 2024-02-01 | 22752 (572.1) | 23804 (527.6) | 4.6 (0.2) | 6.9 (0.7) | 9 | 29 |
| Employer_9 | 2020-06-01 | 2021-06-01 | 2021-07-01 | 2024-01-01 | 178320 (7842) | 198333 (3091.8) | 2 (0.2) | 2.7 (0.3) | 13 | 31 |
| Employer_10 | 2021-09-01 | 2021-12-01 | 2022-01-01 | 2023-08-01 | 11574 (48.9) | 11765 (146.6) | 2.6 (0.2) | 3.2 (0.3) | 4 | 20 |
| Employer_11 | 2022-03-01 | 2023-02-01 | 2023-03-01 | 2024-01-01 | 6455 (307.3) | 7282 (162.4) | 2.6 (0.2) | 4.1 (0.3) | 12 | 11 |
| Employer_12 | 2020-10-01 | 2021-09-01 | 2021-10-01 | 2022-09-01 | 9367 (85.7) | 9136 (113.7) | 4.7 (0.2) | 6.4 (0.8) | 12 | 12 |
| Employer_13 | 2020-08-01 | 2021-07-01 | 2021-08-01 | 2023-09-01 | 8386 (752.9) | 9598 (207.2) | 1.7 (0.1) | 3 (0.5) | 12 | 26 |
| Employer_14 | 2022-05-01 | 2023-04-01 | 2023-05-01 | 2024-04-01 | 78173 (3479.9) | 88735 (1803.4) | 5.1 (0.3) | 6.9 (0.4) | 12 | 12 |
| Employer_15 | 2022-01-01 | 2022-12-01 | 2023-01-01 | 2024-12-01 | 11548 (840.8) | 9339 (1135.6) | 6 (0.2) | 8.8 (0.6) | 12 | 24 |
| Employer_16 | 2020-07-01 | 2021-06-01 | 2021-07-01 | 2024-02-01 | 29921 (1168) | 31644 (415.1) | 2.4 (0.1) | 3.7 (0.4) | 12 | 32 |
| Employer_17 | 2020-07-01 | 2021-06-01 | 2021-07-01 | 2022-06-01 | 13253 (317.6) | 13329 (305.8) | 0.7 (0) | 2.5 (0.3) | 12 | 12 |

**Supplementary Table 1.** *Employer characteristics*. BH = behavioral health.
Employer size and BH utilization are reported as the mean (SD) values across pre-launch and post-launch months.

| | Utilization Rate | | |
|---|---|---|---|
| *Predictors* | *Log-Odds* | *CI* | *p* |
| (Intercept) | -3.62 | -3.86 – -3.38 | **<0.001** |
| phase [Post-launch] | 0.08 | 0.06 – 0.10 | **<0.001** |
| calendar months [1st degree] | 0.56 | 0.51 – 0.60 | **<0.001** |
| calendar months [2nd degree] | 1.04 | 0.94 – 1.14 | **<0.001** |
| calendar months [3rd degree] | 0.70 | 0.63 – 0.76 | **<0.001** |
| program months [1st degree] | -0.06 | -0.08 – -0.04 | **<0.001** |
| program months [2nd degree] | 0.02 | -0.03 – 0.07 | 0.383 |
| program months [3rd degree] | -0.15 | -0.19 – -0.11 | **<0.001** |
| $N_{customer\_name}$ | 17 | | |
| Observations | 545 | | |

**Supplementary Table 2.** *ITS model parameters for behavioral health utilization rate.*

| | **Overall Medical Cost** | | | **Physical Health Cost** | | | **Total BH Cost** | | | **Total Therapy Cost** | | | **Non-Therapy BH Cost** | | |
|---|---|---|---|---|---|---|---|---|---|---|---|---|---|---|---|
| *Predictors* | *Estimates* | *CI* | *p* | *Estimates* | *CI* | *p* | *Estimates* | *CI* | *p* | *Estimates* | *CI* | *p* | *Estimates* | *CI* | *p* |
| (Intercept) | 433.45 | 389.25 – 477.65 | **<0.001** | 390.4 | 352.34 – 428.46 | **<0.001** | 43.47 | 35.56 – 51.37 | **<0.001** | 12.8 | 8.84 – 16.77 | **<0.001** | 30.61 | 25.52 – 35.70 | **<0.001** |
| post launch | -2.16 | -12.92 – 8.61 | 0.694 | -4.54 | -14.59 – 5.50 | 0.375 | 2.4 | 0.74 – 4.07 | **0.005** | 2.82 | 2.18 – 3.46 | **<0.001** | -0.36 | -1.75 – 1.03 | 0.613 |
| calendar months | 4.07 | 2.74 – 5.40 | **<0.001** | 3.63 | 2.39 – 4.86 | **<0.001** | 0.43 | 0.22 – 0.63 | **<0.001** | 0.08 | -0.00 – 0.16 | 0.064 | 0.34 | 0.17 – 0.51 | **<0.001** |
| program months | -2.3 | -3.69 – -0.92 | **0.001** | -2.2 | -3.50 – -0.91 | **0.001** | -0.08 | -0.30 – 0.13 | 0.459 | 0.05 | -0.04 – 0.13 | 0.278 | -0.12 | -0.30 – 0.06 | 0.2 |

**Supplementary Table 3.** *ITS model parameters for overall medical cost, physical health cost, total behavioral health (BH) cost, total therapy cost, and non-therapy behavioral health cost.*
